# An Imaging Risk Score for Bipolar Disorder: Multimodal Comparison and Integration with Polygenic Risk

**DOI:** 10.64898/2026.10.13.26365416

**Authors:** Nadine Parker, Kevin S. O’Connell, Alexey A. Shadrin, Pravesh Parekh, Melody J.Y. Kang, Jakub Kopal, Espen Hagen, Kody G. Kennedy, Yanghee Im, Dag Alnæs, Dylan Hughes, Martin Alda, Nina Alexander, Luisa Altegoer, Ana Aquino-Servín, Jochen Bauer, Dunya Bedir, Francesco Benedetti, Klaus Berger, Michael Berk, Hilary P. Blumberg, Tiana Borgers, Kiymet Bozaoglu, Beatrice Bravi, Geraldo F. Busatto, Erick Jorge Canales, Dara M. Cannon, Xavier Caseras, I-Jou Chi, Paola Fuentes Claramonte, Brett Clementz, Lejla Colic, Emma Corley, Orwa Dandash, Udo Dannlowski, Caroline Demro, Ana M. Díaz-Zuluaga, Karolin Einenkel-Brand, Torbjørn Elvsåshagen, Lisa T. Eyler, Pauline Favre, Kira Flinkenflügel, Maria F. Forte, Janice M. Fullerton, Serena Furno, Gabriel R. Fries, Elliot Gershon, David C. Glahn, Ian H. Gotlib, Roberto Goya-Maldonado, Melissa Green, Dominik Grotegerd, Oliver Gruber, Bartholomeus Haarman, Tim Hahn, Tomas Hajek, Marco Hermesdorf, Josselin Houenou, Fleur M. Howells, Elena I. Ivleva, Hamidreza Jamalabadi, Delfina Janiri, Andreas Jansen, Nooshin Javaheripour, Sarah Keedy, Matthew Kempton, Matcheri Keshavan, Tilo Kircher, Luisa Klahn, Bernd Kraemer, Aodan Laighneach, Mikael Landén, Younghwa Lee, Carlos Lopez-Jaramillo, Walid Makhoul, Ulrik F. Malt, Colm McDonald, Sean McWhinney, Susanne L. Meinert, Philip B. Mitchell, Benson Mwangi, Igor Nenadic, Emilie Olie, Daniele Olivo, Sara Papenbrock, David Parker, Godfrey Pearlson, Fabrizio Piras, Federica Piras, Sara Poletti, Edith Pomarol-Clotet, James Prisciandaro, Yann Quidé, Joaquim Radua, Lisa Rauer, Wiebke Ringels, Elysha Ringin, Gloria Roberts, Amanda L. Rodrigue, Pedro G.P. Rosa, Susan Rossell, Matthew D. Sacchet, Raymond Salvador, Fabio Sambataro, Gabriele Sani, Salvador Sarró, Theodore Satterthwaite, Jonathan Savitz, Freda Scheffler, Maria Serra-Navarro, Yvette I. Sheline, Kang Sim, Jair C. Soares, Márcio Gerhardt Soeiro-de-Souza, Scott Sponheim, Dan J. Stein, Frederike Stein, Benjamin Straube, Carol Tamminga, Henk S. Temmingh, Lea Teutenberg, Florian Thomas-Odenthal, Natasha Topolski, Snezana Urosevic, Paula Usemann, Benedetta Vai, Johanna Valencia-Echeverry, Tamsyn E Van Rheenen, Daniela Vecchio, Eduard Vieta, Enric Vilajosana, Daniel Wolf, Mon-Ju Wu, Albert Yang, Lakshmi Yatham, Marcus V. Zanetti, Giovana B. Zunta-Soares, Carrie E Bearden, Benjamin I. Goldstein, Tobias Kaufmann, Lars T. Westlye, Paul M. Thompson, Anders M. Dale, Christopher R. K. Ching, Ole A. Andreassen

**Affiliations:** Centre for Precision Psychiatry, Division of Mental Health and Addiction, Oslo University Hospital & University of Oslo, Oslo, Norway; KG Jebsen Centre for Neurodevelopmental Disorders, University of Oslo, Oslo, Norway; J. Craig Venter Institute, San Diego, CA USA; Imaging Genetics Center, Mark and Mary Stevens Neuroimaging & Informatics Institute, Keck School of Medicine, University of Southern California; Centre for Addiction and Mental Health, Centre for Youth Bipolar Disorder, Toronto, Canada; Department of Psychology, University of Oslo, Oslo, Norway; University of California, Los Angeles; Los Angeles, CA USA; Department of Psychiatry, Dalhousie University, Canada; Department of Psychiatry and Psychotherapy, University of Marburg, Germany; Center for Mind, Brain and Behavior (CMBB), University of Marburg, Germany; Institute for Translational Psychiatry, University of Münster, Germany; FIDMAG Germanes Hospitalàries Research Foundation, Centro de Investigación Biomédica en Red de Salud Mental (CIBERSAM), Barcelona, Spain; Psychiatry & Clinical Psychobiology, Division of Neuroscience, IRCCS Ospedale San Raffaele, Milan, Italy; Institute of Epidemiology and Social Medicine, University of Münster, Germany; Deakin University, IMPACT Institute – The Institute for Mental and Physical Health and Clinical Translation, School of Medicine, Barwon Health, Geelong, Victoria, Australia; Yale School of Medicine, Department of Psychiatry, New Haven, United States; Murdoch Children’s Research Institute, Parkville Victoria Australia; Laboratory of Psychiatric Neuroimaging (LIM-21), Departamento e Instituto de Psiquiatria, Hospital das Clínicas HCFMUSP, Faculdade de Medicina, Universidade de São Paulo, Brazil; Clinical Neuroimaging Laboratory, Centre for Neuroimaging, Cognition, and Genomics (NICOG), University of Galway, Galway, Ireland; Department of Psychological Medicine and Clinical Neurosciences, School of Medicine, Cardiff University, Cardiff, United Kingdom; Department of Occupational Therapy, Kaohsiung Medical University, Kaohsiung, Taiwan; Biomedical Artificial Intelligence Academy, Kaohsiung Medical University, Kaohsiung, Taiwan; Departament de Psicologia Bàsica, Clínica i Psicobiologia, Universitat Jaume I, Castelló de la Plana (Spain); Departments of Psychology and Neuroscience, BioImaging Research Center, University of Georgia, Athens, Georgia, USA; Jena University Hospital, Department of Psychiatry and Psychotherapy, Jena, Germany; German Center for Mental Health (DZPG), partner site Halle-Jena-Magdeburg Germany; Department of Psychiatry, University of Melbourne, Austin Health, Heidelberg, Victoria Australia; Department of Psychiatry, Medical School and University Medical Center OWL, Protestant Hospital of the Bethel Foundation, Bielefeld University; Center for Intervention and Research on Adaptive and Maladaptive Brain Circuits Underlying Mental Health (C-I-R-C), Site Jena Magdeburg Halle, Germany; Department of Psychology, University of Minnesota, Minneapolis, MN, USA; Department of Psychiatry and Behavioral Sciences, University of Minnesota, Minneapolis, MN, USA; Research Group in Psychiatry, Department of Psychiatry, School of Medicine, Universidad de Antioquia, Medellín, Colombia; Center for Neurobehavioral Genetics, Semel Institute for Neuroscience and Human Behavior, University of California Los Angeles, Los Angeles, USA; Section for Experimental Psychopathology and Neuroimaging, Department of General Psychiatry, Heidelberg University Hospital, Germany; Institute of Clinical Medicine, University of Oslo, Oslo, Norway; Department of Neurology, Oslo University Hospital, Oslo, Norway; Section for Precision Psychiatry, Division of Mental Health and Addiction, Oslo University Hospital, Oslo, Norway, Oslo, Norway; Department of Psychiatry, University of California, San Diego, La Jolla, USA; Université Paris Est Créteil, INSERM Unit 955, Team Translational NeuroPsychiatry, Créteil, France; Neurospin, CEA Paris-Saclay, team UNIACT, Gif-sur-Yvette, France; Institute of Neuroscience, University of Barcelona, Hospital Clinic, IDIBAPS, CIBERSAM, Barcelona, Catalonia, Spain; Swinburne University, Centre for Mental Health and Brain Sciences, Melbourne, Australia; Discipline of Psychiatry and Mental Health, School of Clinical Medicine, Faculty of Medicine and Health, University of New South Wales, Sydney, Australia; Center Of Excellence On Mood Disorders, Louis A. Faillace, MD, Department of Psychiatry and Behavioral Sciences, The University of Texas Health Science Center at Houston, USA; Departments of Psychiatry and Behavioral Neuroscience, University of Chicago, Chicago, IL, USA; Department of Psychiatry, Boston Children’s Hospital and Harvard Medical School, Boston, MA; Department of Psychology, Wu Tsai Neurosciences Institute, Stanford University, Stanford, CA; University Medical Center Goettingen and University Hospital Jena, Department of Psychiatry and Psychotherapy, Jena, Germany; School of Clinical Medicine, University of New South Wales (UNSW), Sydney, Australia; University Medical Center Groningen, the Netherlands; Department of Psychiatry, Dalhousie University, Halifax, Nova Scotia, Canada; Department of Psychiatry, University of Melbourne, Melbourne, Victoria, Australia; Northern Hospital, Mental Health Services, Epping, Victoria, Australia; APHP, Mondor University Hospitals, DMU IMPACT, Créteil, France; Neuroscience Institute, University of Cape Town, Cape Town, South Africa; Department of Psychiatry and Mental Health, University of Cape Town, Cape Town, South Africa; Department of Psychiatry, UT Southwestern Medical Center, 5323 Harry Hines Blvd., Dallas, TX, 75390, USA; Department of Neuroscience, Section of Psychiatry, Università Cattolica del Sacro Cuore, Largo Francesco Vito 1, 00168 Rome, Italy; Department of Psychiatry, Fondazione Policlinico Universitario Agostino Gemelli IRCCS, Largo Agostino Gemelli 8, 00168 Rome, Italy; Department of Psychosis Studies, Institute of Psychiatry, Psychology, and Neuroscience, King’s College London, London United Kingdom; Department of Psychiatry, Beth Israel Deaconess Medical Center, Boston, MA, USA; Department of Psychiatry, Harvard Medical School, Boston, MA, USA; Philipps-Universität Marburg, Department of Psychiatry and Psychotherapy, Marburg, Germany; University of Gothenburg, Institutet of Neuroscience and Physiology, Gothenburg, Sweden; Karolinska Institutet, Department of Medical Epidemiology and Biostatistics, Stockholm, Sweden; Center for Neuromodulation in Depression and Stress, Department of Psychiatry, University of Pennsylvania, Philadelphia, PA, USA; Department of Emergency Psychiatry and Acute Care, Lapeyronie Hospital, CHU Montpellier, Montpellier, France; Department of Neuroscience (DNS), University of Padova, Padova, Italy; Department of Human Genetics, Emory University School of Medicine, Atlanta, Georgia, USA; Psychiatry and Neuroscience, Yale University School of Medicine.; Olin Neuropsychiatry Research Center, and Research Director, Institute of Living, HHC; Neuropsychiatry Laboratory, Department of Clinical Neuroscience and Neurorehabilitation, IRCCS Santa Lucia Foundation, Rome 00179, Italy; Department of Psychiatry and Behavioral Sciences, Medical University of South Carolina, Charleston, SC, USA; NeuroRecovery Research Hub, School of Psychology, University of New South Wales (UNSW) Sydney, Sydney, Australia. 2. Centre for Pain IMPACT, Neuroscience Research Australia, Randwick, Australia; Institut d’Investigacions Biomèdiques August Pi i Sunyer (IDIBAPS), Centro de Investigación Biomédica en Red de Salud Mental (CIBERSAM), Instituto de Salud Carlos III, Barcelona, Spain; Institute of Neuroscience (UBNeuro), University of Barcelona, Spain; University of Melbourne, Department of Psychiatry, Melbourne Medical School, Faculty of Medicine, Dentistry and Health Sciences, Melbourne, Australia; Boston Children’s Hospital, Department of Psychiatry & Behavioral Sciences, Boston, United States; Department of Psychiatry, University of Pennsylvania Perelman School of Medicine, Philadelphia, PA, USA; Laureate Institute for Brain Research, Tulsa, OK, USA; Departments of Psychiatry, Neurology and Radiology Perelman School of Medicine, University of Pennsylvania, Philadelphia, PA, USA; West Region, Institute of Mental Health, Singapore; Yong Loo Lin School of Medicine, National University of Singapore, Singapore; Lee Kong Chian School of Medicine, Nanyang Technological University, Singapore; University of São Paulo, Department and Institute of Psychiatry, São Paulo, Brazil; Mental Health Service Line, Minneapolis VA Health Care System, Minneapolis, MN, USA; SA Medical Research Council Unit on Risk & Resilience in Mental Disorders; Therapeutic and Intermediate Service, Valkenberg Hospital, South Africa; Bipolar and Depressive Disorders Unit, Institute of Neurosciences, Hospital Clinic, Barcelona, Spain; University of Barcelona, Spain; Department of Medical Research, Taipei Veterans General Hospital, Taipei, Taiwan; School of Medicine, National Yang Ming Chiao Tung University, Taipei, Taiwan; Department of Psychiatry, University of British Columbia, Vancouver, British Columbia, Canada; Hospital Sírio-Libanês, São Paulo, Brazil; Department of Psychiatry and Psychotherapy, University of Tübingen, Tübingen, Germany; German Center for Mental Health (DZPG), Germany

## Abstract

**Background:** Biological risk scores provide objective markers of bipolar disorder (BD) liability. Large-scale genome-wide association studies (GWASs) have led to polygenic risk scores (PRSs) that capture genetic liability to BD at the individual level. We lack a comparable large-scale effort to leverage a brain-wide association study (BWAS) to create an imaging risk score (IRS) that captures brain structural liability to BD at the individual-level. Here, we conduct the largest multivariate BWAS of BD and develop a BD IRS. We compare the BD IRS to the PRS and assess their combined predictive performance in multimodal risk score (MRS) models.

**Methods:** Our multivariate BD BWAS used 153 structural neuroimaging features from 3902 cases and 6855 controls (mean age (sd)=35.47 years (13.42), 56.70% female) in a cross-validated machine learning approach. Using the resulting model coefficients, we constructed an IRS. We then evaluated the out-of-sample performance of the IRS, PRS derived from the largest GWAS to date, and combined MRS model using logistic regression in a subset of individuals with imaging and genetic data (n_BD_=486, n_controls_=1701, age (sd)=33.10 (12.02), 56.47% female). We also performed stratified analyses for demographic and BD clinical groups. Prediction performance was quantified using the area under the receiver operating characteristic curve (AUC). Subsequent analyses examined associations between each risk score and BD relevant symptoms in developing youth.

**Outcomes:** In predicting case status, the BD BWAS model achieved a mean AUC_BD_ of 0.62 (sd_BD_=0.08) and in stratified analyses, had higher prediction than subgroup-specific BWAS models. The BD type II BWAS was the only exception (AUC_BDII_=0.60, sd_BDII_=0.07 vs AUC_BD_=0.58, sd_BD_=0.09). The BD PRS had an AUC_BD_ of 0.65 (CI_95%_=0.63-0.68) and a higher AUC than subtype-specific PRS in stratified analyses. The PRS typically attained a higher AUC than the IRS. Notably, the IRS and PRS were uncorrelated (r=0.03, p=0.11). MRS models outperformed the prediction of individual PRS and IRS with the strongest prediction for BD type I (AUC_BD_=0.74, CI_95%_=0.70-0.78). The IRS had the strongest associations with BD-related symptoms in developing youth including increased energy (OR=1.21, p_fdr_=0.03) and distress from psychotic-like experiences (OR=1.23, p_fdr_=0.04).

**Interpretation:** Our BD IRS is a novel biological indicator of BD risk. The lack of correlation between the IRS and PRS along with improved MRS prediction suggests the two risk scores capture some orthogonal liability to BD. We show that integrating imaging and genetics is useful for biological characterization of BD and should be considered for the development of clinically relevant prediction models.

## Introduction

Bipolar disorder (BD) is a heterogenous, episodic, psychiatric disorder with onset typically occurring during adolescence through early adulthood.^1^ The two prominent subtypes of BD are bipolar I disorder (BDI), characterized by depressive and manic episodes, and bipolar II disorder (BDII), characterized by depressive and hypomanic episodes.^1^ Similar to other psychiatric disorders, BD is heritable suggesting there is an underlying biological component that can be used to improve diagnostic and treatment outcomes.^2,3^ However, we lack clear objective biological markers of BD.^4^ Biological risk factors for BD can serve as markers that may inform the characterization of premorbid risk and early detection, quantification of disorder severity, and individualized treatment options.

The fields of psychiatric neuroimaging and genetics have made substantial strides in improving our biological understanding of BD. In psychiatric genetics genome-wide association studies (GWASs) have helped provide insights into the underlying biology of BD revealing relevant genes, mechanisms, cell types, and brain features likely contributing to disorder pathophysiology.^2,5–9^ Neuroimaging provides more proximal biological indicators of BD and its defining symptoms. Similar to a GWAS, a brain-wide association study (BWAS) can be leveraged for biological insights into BD revealing relevant imaging metrics linked to neurobiological features and processes and patterns of BD-related brain alterations linked to patterns of gene expression, biological mechanisms, and cell types.^10–14^ The largest BD BWAS typically use structural features of brain morphology as they are the most readily available and easily combined metrics across cohorts.^10,11,15^ Unlike genetics, imaging metrics are dynamic and are a product of both genetic and environmental factors. A multivariate BWAS associates numerous brain features and an outcome trait often within a cross-validation framework. This approach harnesses instability, gaining reliable associations and prediction performance.^16^ Therefore, GWAS and BWAS provide two biological perspectives that can be leveraged for a greater understanding of BD.

Large-scale GWAS and BWAS of BD have also revealed that while many individual brain measures and genetic variants are associated with the disorder, they have relatively small effect sizes.^10–12,14^ One tactic to capture liability to BD is to aggregate these effects into a single metric. In genetics, the polygenic risk score (PRS) aggregates GWAS effects into a single weighted count of risk alleles. While PRSs for BD have been used in countless research settings, a BD PRS has not achieved clinical utility. There has been increasing interest in developing complementary imaging risk scores (IRSs) for psychiatric disorders which use BWAS effect sizes to generate a weighted metric across imaging features.^17–20^ While there have been previous multivariate BWAS studies of BD,^21,22^ none have generated an IRS and examined its potential for improved prediction performance alongside a BD PRS.

Here we leverage data from an international ENIGMA BD working group collaboration to: (i) perform the largest multivariate BWAS, (ii) create IRSs, (iii) compare the prediction performance of an IRS and PRS, and (iv) evaluate the combined multimodal performance of the IRS and PRS. We also investigate the association between BD symptoms and both the IRS and PRS in developing youth within the Adolescent Brain Cognitive Development℠ Study (ABCD Study^®^). Altogether, we provide a foundation for future studies to use our novel IRS in expanded analyses and illustrate the utility of multimodal biological models for BD.

## Methods

### The ENIGMA Bipolar Disorder Working Group

#### Participants

Across 42 international cohorts, we included 3902 BD cases and 6855 controls with neuroimaging data in the multivariate BWAS [mean age(sd)=35.47(13.42), n_female_=6099 (56.70%)]. Cohort specific demographics, diagnostic instruments, and exclusion criteria are provided in Supplementary Table 1. Institutional review, ethics approval, and participant informed consent was obtained by each participating cohort.

#### MRI Processing

Each cohort applied the standard ENIGMA FreeSurfer processing pipeline to T1-weighted MRI scans along with visual quality control (https://enigma.ini.usc.edu/protocols/imaging-protocols/). Details on cohort-specific image acquisition protocols are in Supplementary Table 1. Pipeline-derived measures of bilateral cortical thickness (n=68) and surface area (n=68) from the Desikan-Kiliany atlas and subcortical brain volumes (n=16) were acquired along with a single measure of total intracranial volume.^23,24^ This resulted in a total of 153 brain features.

### Multivariate Brain-wide Association Study Models

The multivariate BWAS leveraged all 153 structural brain features to predict diagnostic status. We compared five different modelling approaches (support vector machine as well as standard and regularized (elastic net, ridge, and lasso) logistic regression) using a nested cross-validation framework. First, an initial split left one scanner-site out as a validation set. Then, a 5-fold cross-validation procedure was applied to the rest of the sample stratifying sex, diagnostic status, and scanner-sites across folds. Within each fold, the training set was used to pre-process the imaging features by: (i) median imputing missing values, (ii) centering and standardizing features, and (iii) using a mixed-effects model to residualize features for age, age-squared, and sex as fixed effects and scanner-site as a random effect. Pre-processing parameters learned from the training set were applied to the test set. The best performing model across the 5-folds was selected based on the area under the receiver operating characteristic curve (AUC). That model was applied to the held-out validation set and the entire procedure was repeated for each scanner-site (n=53). We selected the best performing modelling approach based on the highest mean AUC across validation sets.

We applied the best performing modelling approach to predict BD status in subgroups restricted to: patients with a BDI diagnosis, patients with a BDII diagnosis, youth (≤21 years), adults (>21 years), females, males, patients with a youth age of onset (diagnosed ≤21 years), patients with an adult age of onset (diagnosed >21 years), and patients with a history of psychosis. For each analysis, we performed the same nested cross-validation procedure described above. To reduce heterogeneity, only scanner-sites with a minimum of 20 cases were included in any prediction analysis. To compare subgroup-specific models to a model trained on the full BD sample, the full BD model was applied to each subgroup-stratified held-out validation set ensuring that the full BD model left out individuals from the validation set to avoid overlap. The BD model (stratified vs full) with the highest mean AUC was considered the best performing.

To extract model coefficients, a final model was run on the full sample. We applied the same pre-processing described above. Extracted coefficients were used for IRS calculations where leave-one-cohort-out runs were performed to avoid target sample overlap. The Supplementary Methods contains details on the BWAS procedure.

### Generation of Imaging Risk Scores

We operationalized the IRS as a weighted mean of the 153 brain structural features. To calculate the IRS for a given cohort, imaging features were first residualized using linear mixed-effects models with age, age-squared and sex as fixed effects and scanner site as a random effect. Coefficients from a given BWAS model were multiplied by the corresponding residualized brain feature. For a given individual, the mean of those weighted brain features was their IRS.

### Generation of Polygenic Risk Scores

We used GWAS summary statistics from the largest GWASs of BD (excluding 23andMe samples) to generate PRS for BD, BDI, and BDII.^5,6^ If a cohort contributed to the GWAS, we generated leave-one-out summary statistics. A consistent protocol was used to generate PRS using imputed genetic data and included: the removal of duplicated variants, genetic ancestry assignment, quantifying genetic relatedness, variant and sample quality control steps, and PRS calculation with the LDpred2-auto method^25^. Details are provided in the Supplementary Methods with all scripts and instructions for generating the PRS provided on GitHub (https://github.com/nadineparker/ENIGMA_BD_PRS).

### Risk Score Prediction Analyses

Six clinical cohorts provided both imaging and genetic data amounting to an unrelated sample of 573 cases and 1762 controls. To reduce heterogeneity for a given prediction analysis, we selected individuals of European genetic ancestry which comprised 94% of the sample and only included cohorts with at least 20 cases (n_BD_=489, n_controls_=1704). All analyses were conducted in the combined sample with post-hoc analyses conducted in each individual cohort. We additionally performed post-hoc prediction analyses using the next largest subset of participants which were of East Asian genetic ancestry (n_BD_=37, n_controls_=16). For the distribution of genetic ancestry within cohorts see Supplementary Table 1.

We used logistic regression to assess PRS prediction of BD, BDI, and BDII compared to controls quantifying performance using the AUC. To facilitate comparison with the IRS, PRSs were residualized for age, sex, and the first 20 genetic principal components. We used the BD PRS to predict case status in both the BDI and BDII subgroups and compare AUC values with subtype-specific PRSs.

We contrast as well as combine the prediction performance of the PRS and IRS using logistic regression. PRS-only, IRS-only, and multimodal risk score (MRS: both PRS and IRS in logistic regression) models were used to predict BD case status in full and subgroup-stratified analyses (see above). The Pearson correlation coefficient between the PRS and IRS was estimated in the full and stratified analyses. We used logistic regression to individually test for differences in PRS and IRS among patients taking vs not taking a given medication category at the time of scanning (i.e., lithium, antiepileptics, generation one antipsychotics, generation two antipsychotics, and antidepressants).

### Risk Score Associations with BD Symptoms in Youth

To examine associations between BD-related symptoms and each risk score, we leveraged the ABCD Study**^®^** which follows nearly 12,000 youth initially recruited from 21 sites across the United States at age 9-10 years. Here, we used data from release 7.0.^26^ Details on the imaging protocol have previously been described.^27,28^ To calculate the BD IRS, imaging features were extracted from baseline and follow-up visits at year 2, 4, and 6. Genotyping and imputation details have also been previously described.^29,30^ We used released imputed genotypes to calculate the PRS in the same manner as described above.

We leveraged summary scores for BD-relevant symptoms from three questionnaires. From the Kiddie Schedule for Affective Disorders and Schizophrenia BD questionnaire, we used all available summary scores for a past experience of symptoms of BD retaining responses from both a parent and the youth.^31^ From the 7-Up Mania questionnaire, we binarized the sum score such that youth reporting the experience any manic symptoms (sum score >0) comprised one group and those with no symptoms (sum score=0) were in the other.^32^ From the Prodromal Psychosis Scale completed by the youth only, we used summary scores for current and persistent distress.^33,34^ Additionally, we binarized the count for endorsing (i) a symptom and (ii) being bothered by a symptom (never=0 vs ever>=1). All questionnaire data were obtained at the same visit as the neuroimaging except the 7-Up Mania data. Here, assessments were acquired at follow-up visits 1, 3, 5, and 7 which were then aligned to the preceding imaging visit (baseline, and years 2, 4, 6). To assess the association between the PRS or IRS with the endorsement of symptoms, we used logistic regression including all participants in this diverse, population-based sample of developing youth. The same procedures for residualization of covariates described above were applied and age was measured in months. Post-hoc analyses were performed within genetic-ancestry subgroups.

### Software and Statistics

All analyses in this study were performed using R v4.4.1 or Python v3.14. To test for a difference in AUC between models, we used bootstrapping. The Benjamini Hochberg method was used to correct for multiple comparisons for each set of analyses.

## Results

### Multivariate Brain-wide Association Study of Bipolar Disorder

A total of 3902 cases and 6855 controls were leveraged for the multivariate BWAS of BD [mean age(sd)=35.47(13.42), n_female_=6099 (56.70%)]. Across the modelling approaches, ridge regression achieved the highest mean AUC (AUC=0.62, sd=0.08; Supplementary Table 2). In stratified analyses, the model trained on the full BD sample achieved a higher mean AUC than subgroup specific models (Figure 1A). The only exception was the differentiation of BDII cases from controls (total n_cases_=581, n_controls_=1282; BDII: mean AUC=0.60, sd=0.07; BD: mean AUC=0.58, sd=0.09). Therefore, the BD model was used to generate all IRS except for analyses differentiating BDII cases from controls where the BDII model was used. The coefficients of the BD and BDII models had a similar spatial distribution across the brain with subtle differences in magnitude (r=0.91, p=7.63e-59; Figure 1B). All model coefficients are in Supplementary Table 3.

**Figure 1.**
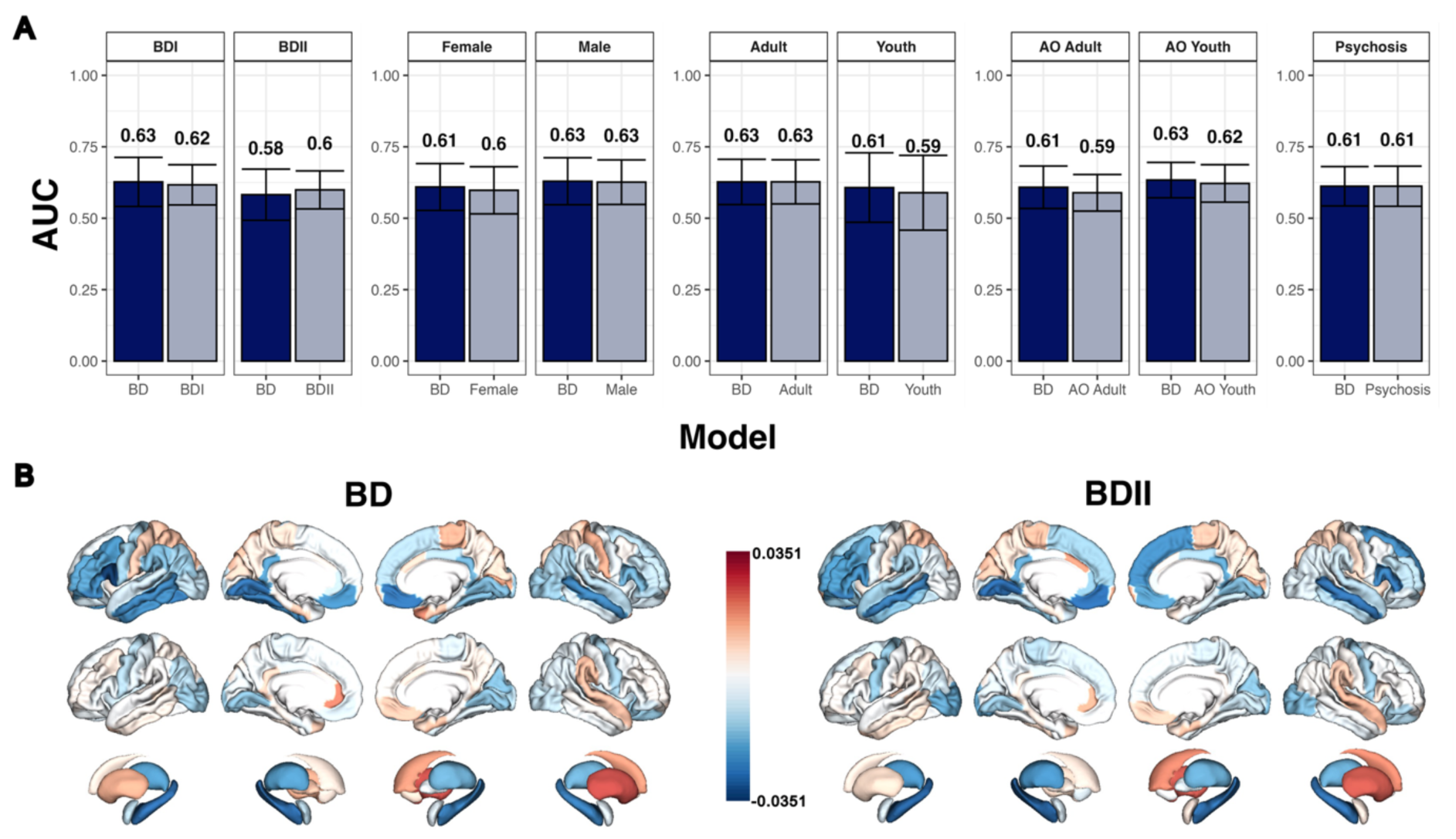
Multivariate Brain-Wide Association Study Model Comparisons and Coefficients. A) Model performance for subgroups of bipolar disorder (BD) cases compared to controls. Prediction performance is measured using the area under the receiver operating curve (AUC). Each bar represents the mean AUC across iterations with error bars representing one standard deviation. The performance of the full BD model (dark blue) is contrasted with the subgroup-specific model (light blue) for each subgroup. B) The full BD and BD type II (BDII) model coefficients for cortical thickness (top), cortical surface area (middle), and subcortical volumes (bottom). Red indicates larger values in BD cases vs controls while blue indicates smaller values. AO=Age of onset where adults are defined as greater than 21 years old.

### Performance of Polygenic Risk Scores

Genetic data from 2187 unrelated participants (BD=486, controls=1701) of European ancestry were included in PRS analyses. The participants had a mean age of 33.10 years (sd=12.02) and 1235 (56.47%) were female. In this sample, the BD PRS derived from a multi-ancestry GWAS attained a higher AUC than that derived from a European ancestry GWAS (Supplementary Table 4). Although this difference was not significant (Supplementary Table 5), we proceed with the multi-ancestry-derived PRS for all further analyses. Prediction performances for the BD, BDI, and BDII PRS are depicted in (Figure 2). The BD PRS achieved an AUC of 0.65 in the combined sample with a range across cohorts from 0.56 in a youth sample to 0.72 in an adult sample. In stratified analyses, the BD PRS had a higher AUC in differentiating BDI cases (n_total_=200) from controls (n_total_=1655) and BDII cases (n_total_=79) from controls (n_total_=1655) than subtype-specific PRS (Figure 2, Supplementary Table 4) but only the latter was statistically significant (Supplementary Table 5). The BD PRS had a higher AUC in the stratified analyses. However, the youth sample with the lowest BD PRS performance did not contribute to the subtype stratified analyses due to low sample size. In analyses of participants of east Asian genetic ancestry (n=53; mean age=34, sd=11.18; n=32 females) the AUC for the PRS derived from the European ancestry GWAS was 0.51 which significantly increased to 0.59 when using the multi-ancestry GWAS (p=0.02).

**Figure 2.**
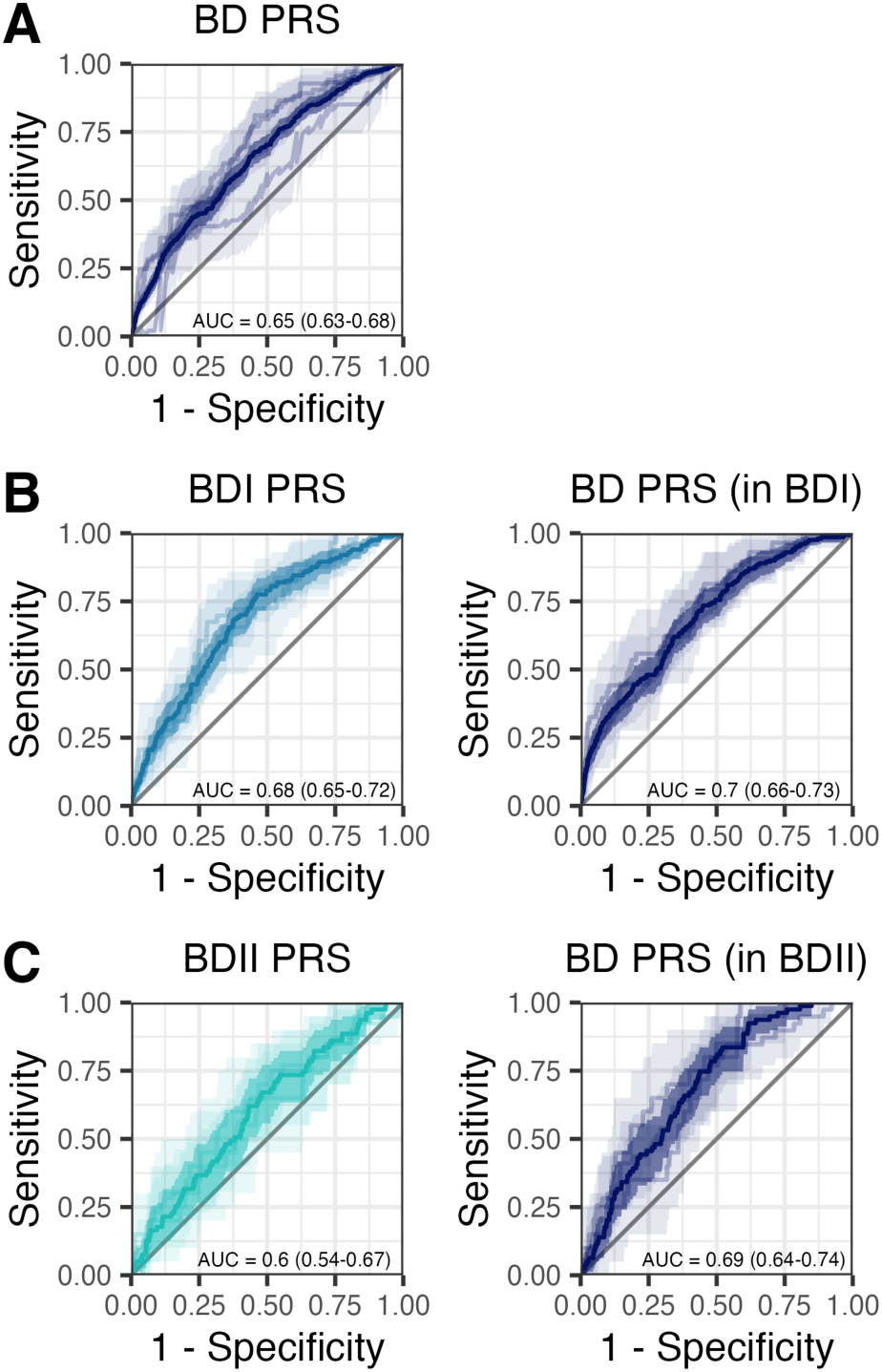
Polygenic risk score prediction performance. Receiver operating characteristic curves for prediction performance in A) BD cases vs controls, B) BDI cases vs controls, and C) BDII cases vs controls. The shaded area represents the 95% confidence interval. The combined sampled is depicted by the dark line and shading. Each individual cohort is depicted by the lighter lines and shading. The area under the receiver operating characteristic curve (AUC) along with the 95% confidence interval is displayed in the bottom right corner for the combined sample.

### Multimodal Risk Score Models Improve Prediction

Analyses comparing the PRS, IRS, and MRS models were conducted in the same sample of 2187 participants with genetic and imaging data. The BD PRS was not correlated with the IRS in the combined sample (r=0.03, p=0.11) and only one cohort had a weak positive correlation (r=0.06, p=0.04; Supplementary Table 6). In differentiating BD cases from controls in the full and stratified analyses, the PRS consistently achieved a higher AUC than the IRS except for the youth stratified analyses (Supplementary Table 7). In youth, only the IRS, not the PRS, significantly differentiated BD cases from controls in the combined sample (OR=1.48, p_fdr_=7.74e-3; Supplementary Table 8). Compared to the IRS and PRS only models, the MRS models achieved the largest prediction AUC (Figure 3, Supplementary Table 7) with prediction performance significantly greater than the leading individual risk score for all analyses except the BDII and youth stratified analyses (Supplementary Table 9). The strongest prediction performance resulted in an AUC of 0.74 for the MRS model differentiating BDI cases from controls in the combined sample. Here, both PRS and IRS were significantly associated with BD status (PRS: OR=2.19, p_fdr_=1.51e-19; IRS: OR=1.64, p_fdr_=1.64e-9; Supplementary Table 8). Analyses of participants of East Asian genetic ancestry showed the PRS and IRS were correlated (r=0.44, p=1.13e-3) but neither significantly differentiated cases from controls (PRS: OR=1.48, p=0.20, AUC=0.59; IRS: OR=1.63, p=0.15, AUC=0.63).

**Figure 3.**
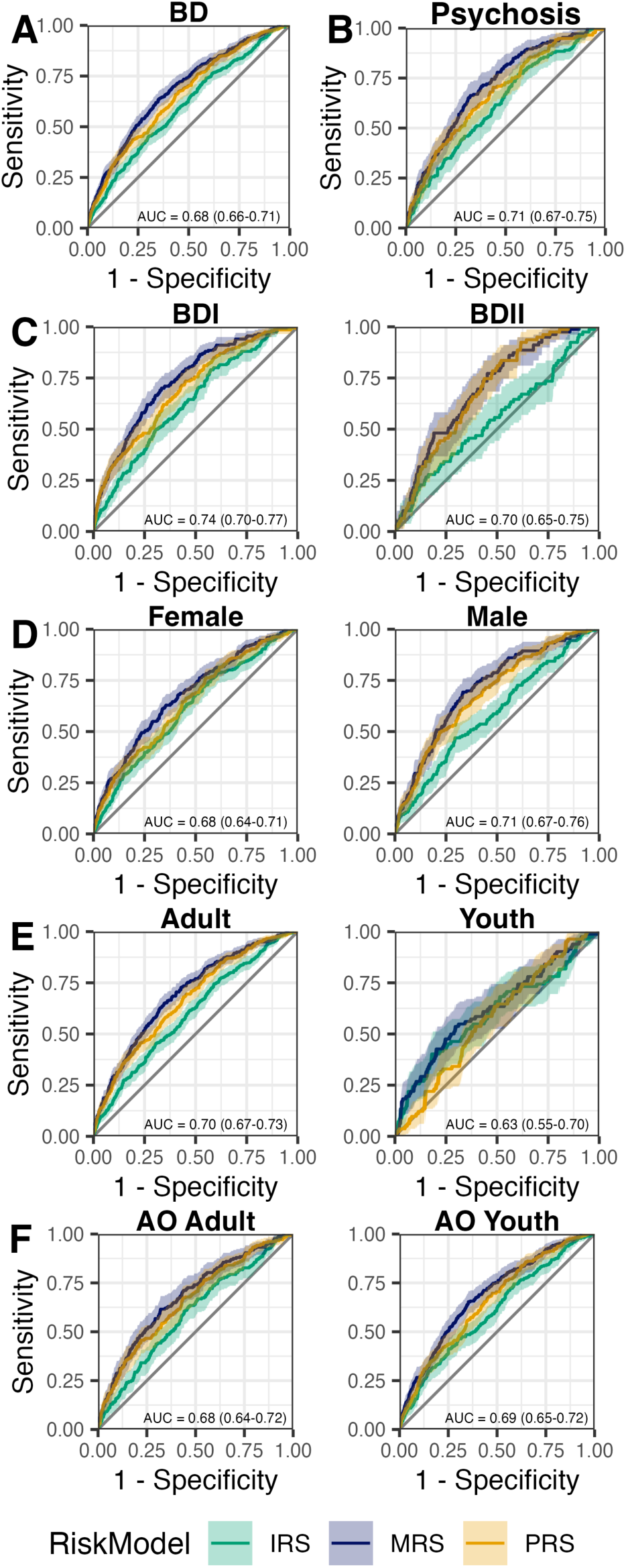
Polygenic, Imaging, and Multimodal Risk Score Model Prediction Performance. Receiver operating characteristic curves for cases vs controls in the full BD sample (A) and stratified samples (B-F). The shaded area represents the 95% confidence interval. The area under the receiver operating characteristic curve (AUC) along with the 95% confidence interval is displayed in the bottom right corner for the multimodal risk score model in the combined sample across contributing cohorts.

In analyses comparing patients taking different medications, a nominally larger PRS was associated with lithium usage (beta=0.13, p_fdr_=0.19). Similarly, a greater BD and BDII IRS was nominally associated with generation one antipsychotic use (BD IRS: beta=8.18e-04, p_fdr_=0.07; BDII IRS: beta=6.44e-04, p_fdr_=0.07). However, none of these associations survived correction for multiple comparisons (Supplementary Table 10).

### Risk Score Associations with Symptoms in Youth

In a total of 9536 (n_female_=4521, 47.41%) unrelated youth ranging in age from 8.32 to 18.00 years across four time-points, we observed associations between risk scores and BD-related symptoms. The IRS was significantly associated with parent reports of increased energy (OR=1.21, p_fdr_=0.03) and youth reports of greater currently distressing psychotic-like experiences (OR=1.23, p_fdr_=0.04; Figure 4). Only these associations survived correction for multiple comparisons (Supplementary Table 11). Additional analyses in genetic ancestry-specific subgroups revealed the strongest associations in the admixed American subgroup between the IRS and less racing thoughts as well as the PRS and greater pressured speech (Supplementary Table 12).

**Figure 4.**
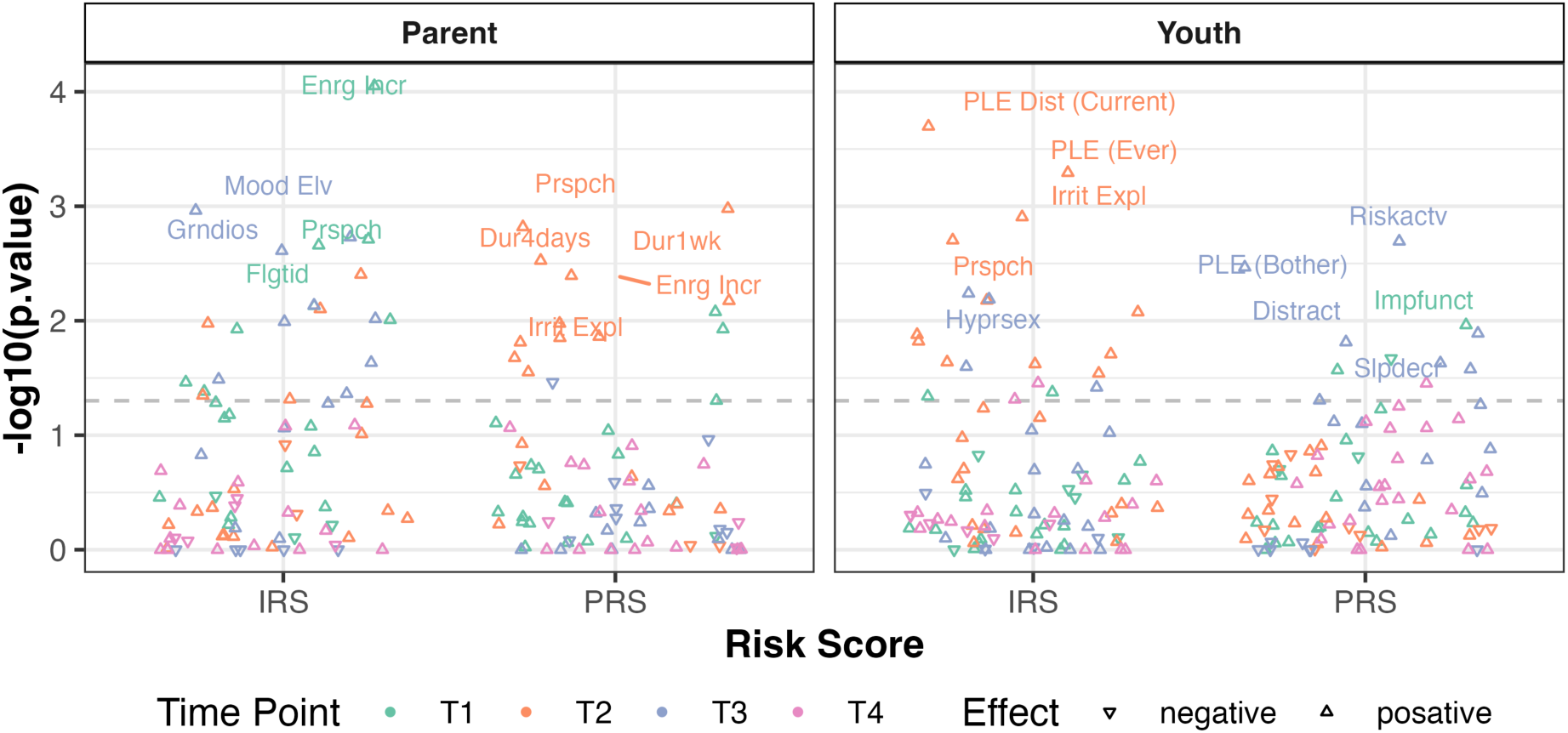
Risk Score Associations with Bipolar Disorder Relevant Symptoms in Developing Youth. The association between the dipolar disorder (BD) IRS and PRS with the presence of different bipolar disorder related symptoms are depicted. The colors represent the four imaging acquisition time-points. The left plot represents associations with symptomology as reported by a parent. The right plot represents associations with symptomology as self-reported by the youth. The horizontal dotted line represents the nominal significance threshold (p=0.05). EnrgIncr: Increased Energy; Mood Elv: Elevated Mood; Grndios: Grandiosity; Prspch: Pressured Speech; Flgtid: Flight of Ideas; Dur4days: BD duration lasting 4 days; Dur1wk: BD duration lasting 1 week; Irrit Expl: Explosive Irritability; PLE Dist (Current): Psychotic-Like Experiences Current Distress; PLE (Ever): Ever Having a Psychotic-Like Experience; Hyprsex: Hypersexuality; Riskactv: Excessive Involvement in High-risk Activities; PLE (Bother): Ever Bothered by a Psychotic-Like Experience; Impfunct: Impairment in Function due to BD; Slpdecr: Decreased Need for Sleep.

## Discussion

In this study we performed the largest multivariate BWAS of BD to derive an IRS. We subsequently compared and combined the IRS with a BD PRS. We found that the BD BWAS model trained on the full sample achieved better prediction performance in subgroup-stratified analyses except for the BDII group. Similarly, the BD PRS outperformed subtype-specific PRS in their respective stratified models. In the full and most stratified analyses, the combined prediction performance of the MRS model was greater than the leading individual risk score. When comparing the two risk scores, the BD PRS tended to achieve higher AUC values than the IRS. Finally, in a developing youth sample, the IRS exhibited the strongest associations with BD symptomology. The combined findings illustrate the utility of the IRS and its inclusion in MRS models that leverage biological indicators for the prediction and characterization of BD.

No previous study has generated and characterized an IRS for BD derived from such a large-scale multivariate BWAS. Here our full BWAS model reached a mean AUC of 0.68 which is higher than a previous BWAS of BD that yielded an AUC of 0.61 using a similar leave-one-site-out approach.^21^ However, the same study using a different individual-level analytical approach did achieve an AUC of 0.71.^21^ Further advances in BWAS modeling and larger sample sizes may result in greater prediction accuracy. Our novel approach provides a simple translation of our large-scale BWAS to an individual-level IRS that is easily applicable to any imaging cohort.

Currently, power outperforms specificity. We show that the BD PRS outperformed the BDI and BDII PRSs in subgroup stratified analyses. Moreover, the BWAS that leveraged the full sample, and not specific subgroups, typically performed best in stratified analyses. The BDII IRS was the only exception suggesting the pattern of structural brain alterations in these patients differs enough to be detectable with the current sample size. With even larger samples more subgroup-specific risk scores may follow the same trend. This study provides a novel comparison of a PRS and IRS for BD where we find the former tends to outperform the latter. This too may reflect a difference in power where the PRS is derived from a multi-ancestry GWAS that included 67,948 cases and 867,710 controls, a sample size orders of magnitude larger than our BWAS used to derive the IRS.^5^ However, our association analyses in a large developing youth sample suggests the IRS may have greater utility in this subgroup. A previous study leveraging an IRS approach in youth samples observed associations with general psychopathology.^18^ These IRS associations in youth samples suggest the potential utility of the IRS for early detection of BD risk. More work is required to expand our understanding of the optimal clinical setting and utility of each risk score.

The finding that the IRS and PRS were uncorrelated and their combination in MRS models improved prediction, suggests the two risk scores capture some orthogonal liability to BD. There are several putative explanations. First, the IRS can capture disorder biology not currently captured by the PRS. That is, the current largest GWAS for BD is not saturated, meaning there are still many genetic associations yet to be discovered.^5^ Therefore, the IRS may capture this currently undiscovered, genetically-regulated biology. Second, even when the GWAS for BD achieves saturation, it will only capture linear genetic associations for common genetic variants. The contribution of rare genetic variants, complex non-linear genetic interactions, and higher-level biological interactions only partially, or not at all, informed by genetics will not be captured in the saturated GWAS for BD or any subsequently derived PRS. Therefore, the BD PRS is unlikely to capture all relevant disorder biology and can be supplemented by an IRS and other biological risk scores. Finally, the IRS can capture non-genetically regulated or environmental effects on the brain. Internal experiences and environmental exposures that can alter brain structure may differ between patients and controls.^35,36^ While some of these exposures are potential confounders, others are important disorder-related characteristics that are better captured by dynamic brain imaging features than static genetic effects. Therefore, this study shows that combining neuroimaging and genetic risk for BD is both warranted and useful.

There are some study limitations to consider. We only use structural neuroimaging features in this study. While this helps maintain large sample sizes, future studies extending our framework to include additional imaging modalities are needed. Medication treatment of patients with BD may result in alterations in brain structure that are captured by the IRS. This may bias prediction performance and potentially contribute to the lack of correlation between the IRS and PRS. However, we show that patients taking different classes of medications had subtle or no differences in their IRS or PRS. Our prediction analyses were predominantly in participants of European genetic ancestry. While some analyses were performed in a small sample of East Asian genetic ancestry, future work should further assess the prediction performance of the IRS and MRS models in more diverse clinical samples.

Biological risk scores for BD can serve as objective measures that aid in early or premorbid risk detection, characterization of disorder severity, and treatment guidance. While current prediction performance of PRS and our IRS for BD may not reach clinical thresholds, our MRS models show the utility of combining biological indicators for improved prediction. Further work is needed to explore how these risk scores and multimodal models that incorporate biological, clinical, and additional variables could inform clinical decision-making. Here, we lay a foundation for a multimodal risk scoring framework for BD that can potentially be expanded and implemented in clinical settings.

## Data Availability

BWAS model coefficients for generating the IRS are available in the supplementary materials. The BD GWAS summary statistics used for generating the PRS are available from the Psychiatric Genomics Consortium (https://pgc.unc.edu/for-researchers/download-results/). The scripts used to generate the PRS are available on GitHub (https://github.com/nadineparker/ENIGMA_BD_PRS).

https://github.com/nadineparker/ENIGMA_BD_PRS

## Contributions

Conceptualization: NP, KSO, AA, PP, AMD, OAA; Data curation - original: all listed authors; Data curation – finalized: NP, KSO, MJYK, YI, CRKC; Analytical Software and Infrastructure: NP, AA, PP, EH, AMD; Formal analyses: NP, AA; Visualization: NP, JK; Writing – original draft: NP; Writing – review & editing: all listed authors.

## Acknowledgments

Data used in the preparation of this article were obtained from the Adolescent Brain Cognitive Development™ (ABCD) Study, held in the NIH Brain Development Cohorts Data Sharing Platform. This is a multisite, longitudinal study designed to recruit more than 10,000 children aged 9-10 and follow them over 10 years into early adulthood. The ABCD Study® is supported by the National Institutes of Health and additional federal partners under award numbers: U01DA041048, U01DA050989, U01DA051016, U01DA041022, U01DA051018, U01DA051037, U01DA050987, U01DA041174, U01DA041106, U01DA041117, U01DA041028, U01DA041134, U01DA050988, U01DA051039, U01DA041156, U01DA041025, U01DA041120, U01DA051038, U01DA041148, U01DA041093, U01DA041089, U24DA041123, U24DA041147. A full list of supporters is available at Federal Partners - ABCD Study. ABCD Consortium investigators designed and implemented the study and/or provided data but did not necessarily participate in the analysis or writing of this report. This manuscript reflects the views of the authors and may not reflect the opinions or views of the NIH or ABCD Consortium investigators. The ABCD data repository grows and changes over time. The ABCD data used in this report came from NBDC Digital Object Identifier (<u>ABCD RELEASE 7.0 (DOI 10.1.66.93/8F3W-5260</u>)

BG was supported by the Canadian Institutes of Health Research (MOP 136947, PJT 162110, MOP 119525) and a Miner’s Lamp Innovation Fund from the University of Toronto. The BiDirect Study was supported by grants of the German Ministry of Research and Education (BMBF) to the University of Münster (01ER0816 and 01ER1506). MB is supported by a NHMRC Leadership 3 Investigator grant (GNT2017131). HPB was supported by the National Institute on Drug Abuse RL1DA024856, National Institute of Mental Health RC1MH088366, R01MH69747, R01MH070902, R01MH113230, R61MH111929 and the National Center for Advancing Translational Science UL1TR000142. The Sao Paulo (Brazil) studies have been supported by grants from FAPESP-Brazil (2013/03905-4), CNPq-Brazil (#478466/2009 & 480370/2009), and the Brain & Behavior Research Foundation (2010 NARSAD Independent Investigator Award granted to Geraldo F. Busatto). DMC is supported by the Health Research Board Ireland (HRA-POR-324). LC was supported by the Interdisciplinary Center of Clinical Research of the Medical Faculty Jena (AMS-21), and the Federal Ministry for Research, Technology and Space (01EE2507F). UD was funded by the German Research Foundation (DFG, grant FOR2107 DA1151/5-1, DA1151/5-2, DA1151/9-1, DA1151/10-1, DA1151/11-1; DA1151/21-1 to UD; SFB/TRR 393, project grant no 521379614) and the Interdisciplinary Center for Clinical Research (IZKF) of the medical faculty of Münster (grant Dan3/016/26). LTE was supported by MH083968. MFF received the support of “Contratos predoctorales de formación en investigación en salud” (PFIS22) (FI22/00185) from the Instituto de Salud Carlos III (ISCIII) with European funds from the Recovery, Transformation and Resilience Plan, by virtue of the Resolution of the Directorate of the Carlos III Health Institute, O.A., M.P. of December 14, 2022, granting Predoctoral Research Training Contracts in Health (PFIS Contracts). Funded by the European Union Next Generation EU. She has also been granted by M-AES mobility fellowship (MV24/00050). DCG is supported by MH077945 MH106324 NIH M01 RR001346 NIH/NIMH R01 MH080912. IHG is supported by R37MH101495. The Imaging Genetics in Psychosis (IGP) study was supported by Project Grants from the Australian National Health and Medical research Council (NHMRC; APP630471 and APP1081603). Melissa J. Green was supported by an Australian Research Council Future Fellowship (FT0991511) and a R.D. Wright Biomedical Career Development Award from the NHMRC (1061875). The FOR2107 cohorts (Marburg and Münster) and the Münster Neuroimaging Cohort are part of the German multicenter consortium “Neurobiology of Affective Disorders. A translational perspective on brain structure and function“, funded by the German Research Foundation (Deutsche Forschungsgemeinschaft DFG; Forschungsgruppe/Research Unit FOR2107). Principal investigators (PIs) with respective areas of responsibility in the FOR2107 consortium are: Work Package WP1, FOR2107/MACS cohort and brain imaging: Tilo Kircher (speaker FOR2107; DFG grant numbers KI588/14-1, KI588/14-2, KI588/20-1, KI588/22-1), Udo Dannlowski (co-speaker FOR2107; DA 1151/5-1, DA 1151/5-2, DA1151/6-1, DA1151/9-1, DA1151/10-1, DA1151/11-1). JH was supported by Agence Nationale de la Recherche (Grant Nos. ANR-11-IDEX-0004 Labex BioPsy and ANR-10-COHO-10-01 psyCOH), Fondation pour la Recherche Médicale (Bioinformatique pour la biologie 2014), and Fondation de l’Avenir. ALK was supported by the Märta Lundqvist’s Foundation and the Kurt och Ingrid Dahréns Foundation. AL is supported by Health Research Board Ireland (HRA-POR-324). The St. Göran study (SBP) was supported by grants from the Swedish Research Council (2022-01643), the Swedish foundation for Strategic Research (KF10-0039), Hjärnfonden/the Swedish Brain foundation (FO2025-0004-HK-212), and the Swedish Government under the LUA/ALF agreement (ALFGBG-1005343). The Medellin collections were funded by the PRISMA UNION TEMPORAL (MinCiencias, Colombia), and CODI-UdeA. The UNSW Sydney “Kids and Sibs” Study was supported by the Australian National Medical and Health Research Council (Program Grant 1037196 and Investigator Grant 1177991 to Philip B. Mitchell). The MUSC site was funded by the National Institute on Alcohol Abuse and Alcoholism (grant K23 AA020842 to James J. Prisciandaro), with additional support provided by a Brain and Behavior Research Young Investigator Grant. The Imaging Genetics in Psychosis (IGP) study was supported by Project Grants from the Australian National Health and Medical research Council (NHMRC; APP630471 and APP1081603). FS is supported by Medical Research Council (SAMRC) of South Africa through its Division of Research Capacity Development under the Early Investigators Programme from funding received from the South African National Treasury. The content and findings reported/ illustrated are the sole deduction, view and responsibility of the researcher and do not reflect the official position and sentiments of the SAMRC. KS was supported by a research grant from the Singapore Bioimaging Consortium (RP C009/2006). JCS was supported in part by NIMH grant R01 MH085667, the Dunn Foundation and the Pat Rutherford, Jr. Endowed Chair in Psychiatry. DJS was supported by the SAMRC. TVR was supported by a Al and Val Rosenstrauss fellowship form the Rebecca L Cooper Medical Research Foundation. The COGSBD study was financially supported by the NHMRC (1060664), Henry Freeman Trust, Jack Brockhoff Foundation, University of Melbourne, Barbara Dicker Brain Sciences Foundation, Rebecca L Cooper Foundation and the Society of Mental Health Research. The authors acknowledge the facilities and scientific assistance of the National Imaging Facility, a National Collaborative Research Infrastructure Strategy (NCRIS) capability, at the Swinburne Neuroimaging Facility, Swinburne University of Technology. CEB is supported by NIH/NIMH R01MH127165, NIH/NINDS/NIMH T32NS048004, R01MH129858, R01MH123157. OAA was supported by the Research Council of Norway (#324499), NordForsk (#164218), NIMH (1R01MH129742), European Union’s Horizon2020 Research and Innovation Action Grant (# 847776 CoMorMent) and the Kristian Gerhard Jebsen Stiftelsen (SKGJ-MED-021). The National Institutes of Health (U01MH108150 to Scott R. Sponheim and K01MH093621 to Snezana S. Urosevic) and the U.S. Department of Veterans Affairs (I01CX000227 to Scott R. Sponheim).

