## Supplementary Methods for "An Imaging Risk Score for Bipolar Disorder: Multimodal Comparison and Integration with Polygenic Risk"

### Description of the Multivariate BWAS Models

We conducted a multivariate BWAS using all 157 structural brain features to predict BD status. Any participants missing more than 50% of the imaging features was removed from analyses. Initially we compared the prediction performance of five different regression modelling approaches: (support vector machine as well as standard and regularized (elastic net, ridge, and lasso) logistic regression) using a nested cross-validation framework. First, the full sample was split such that one scanning site was held-out as a final validation set. Within the larger multi-cohort sample, a 5-fold cross validation procedure was performed stratifying by sex, diagnostic status, and cohort, such that each fold maintained a consistent number of each category. Within each fold, the training set containing 80% of the sample was used to process the imaging features by: (i) using the median to impute missing values, (ii) centering and scaling all features, and (iii) using a mixed effects model to residualize each feature for age, age-squared, and sex as fixed effects and scanner site as a random effect. The processing parameters from the training data were then applied to the test set. Then for a given modelling approach (e.g., lasso) 5-fold cross-validation was conducted where parameters are tuned (see below). The held-out test set performance was used to select the model with the best prediction performance operationalized as the highest area under the receiver operating characteristic curve (AUC). That best performing model was applied to both the full train (all data that went into cross-validation) and the final held-out validation set comprised of a single imaging site. Before doing so, data from the full train set was processed in the same manner as previously described (i.e., median imputation, centering and scaling and residualization with a mixed-effect model). Those parameters were applied to the final held-out validation set. The entire procedure was repeated over each imaging site (n=53) and the performance of each of the five modelling approaches was compared. The approach with the highest mean AUC value was selected for further analyses.

Once the modelling approached was selected, we performed stratified analyses predicting BD status. The following subgroup analyses were performed restricting to: patients with a BDI diagnosis, patients with a BDII diagnosis, youth (participants aged 21 and under), adults (participants older than 21), females, males, patients with a youth age of onset (diagnosed at age 21 or younger), patients with an adult age of onset (diagnosed over the age of 21), and BD patients with a history of psychosis. In each stratified analysis the same nested cross-validation procedure was performed using the selected modelling approach. To reduce heterogeneity due to limited sample size, only imaging sites with a minimum of 20 cases were included in any given stratified analysis. As described above the final prediction performance was operationalized as the mean AUC across all held-out validation sets. We also compared if subgroup-specific models outperformed models trained on the full BD sample. To do so, the full BD model was applied to each subgroup stratified held-out validation set. Importantly, the full BD model applied did not include the imaging site used as the held-out validation set to avoid sample overlap. The BD model (stratified vs full) with the highest mean AUC was considered the best performing within each stratified analysis.

Model coefficients for the imaging features were extracted from a final run of the best performing models which was applied to the full sample. Before running the final model, the data was processed as described above where individuals with >50% of features missing are removed from analyses and median imputation, centering and scaling and residualization with mixed-effect models. The extracted coefficients were used for subsequent IRS calculations. Importantly, leave-one-site-out runs were also performed such that IRS can be calculated for a given site without sample overlap.

All analyses were conducted within R v4.4.1. Logistic regression models were run using the standard stats package. Linear support vector machine models were run using the kernlab package where the C parameter was held at the default value of 1. Ridge and lasso models were run using glmnet where alpha was set to 0 and 1 respectively and lambda was tuned within cross-validation. Elastic net models were run using glmnet within the caret package such that both alpha and lambda were tuned using default settings.

### Description of the Polygenic Risk Score Procedure

Information on genotype array, imputation software and reference genome, as well as post imputation quality filtering for sites that provided genetic data can be found in supplementary table 1. All imputed genotypes were then processed using our standardized polygenic risk score (PRS) protocol outlined on the project GitHub page (https://github.com/nadineparker/ENIGMA_BD_PRS). All necessary software were included in Singularity of Docker containers. The protocol uses genomic references and summary statistics in genomic build GRCh38. If the imputed genotypes were in a different build, they were lifted to build GRCh38 using the LiftOver tool and a standardized script also provided on the above linked GitHub page. All necessary files (summary statistics, genomic references, containers, etc.) too large for the project GitHub page were stored on a project figshare website (https://doi.org/10.6084/m9.figshare.25872160).

The PRS protocol first removes any duplicated genetic variants and then performs genetic ancestry assignment using the superpopulations from the 1000 genomes genomic reference. The ancestry assignment procedure first performs some quality control procedures that identify and restrict 1000 genome variants to those common across ancestries, aligns alleles in the target cohort to the genomic reference, performs strand flipping when necessary, performs pruning to select independent variants all using Plink v2.0.[1] Then the target cohort genetic principal components (PCs) are compared to those of the 3202 samples from the 1000 genomes phase 3 dataset.[2] The 1KGP samples had previously been assigned to one of five continental ancestry groups: African (AFR, n=893), Admixed American (AMR, n=490), East Asian (EAS, n=585), European (EUR, n=633), and South Asian (SAS, n=601). Reference data were obtained from https://www.cog-genomics.org/plink/2.0/resources#phase3_1kg. Before calculating PCs, the 3202

1KGP samples were first pruned to retain 2573 unrelated samples up to the second degree of relatedness (--king-cutoff 0.0884). PCs were calculated using these unrelated samples. PCs for all 1KGP samples and for samples in the target dataset were then obtained by projection onto the PC basis derived from the unrelated 1KGP samples. A histogram-based gradient boosting classifier (HistGradientBoostingClassifier) with default options, implemented in the python package scikit-learn was trained using the first four PCs of the 1KGP samples and their predefined ancestry-group labels. The trained classifier was subsequently applied to infer the genetic ancestry of samples in the target dataset. Afterwards, we also calculated genetic relatedness in the target sample using the pruned genotypes to capture first and second degree relatives.

After ancestry assignment, quality control of the genetic data is performed within genetic ancestries to avoid removal of individuals within minority populations that may appear as outliers in the combined sample. Plink v1.9 is used to generate ancestry specific bed files, apply a minor allele frequency threshold of 0.01, do Hardy Weinberg equilibrium thresholding at 1e10, filter variants based on a missingness threshold of 0.1, and perform heterozygosity filtering such that individuals with an F-value greater than 4 standard deviations from the mean were removed. This quality control procedure was slightly augmented if the number of samples with a given ancestry dropped below 10. Here, due to low sample size, the minor allele frequency filtering was dropped, the genotype missingness filter was raised for 0.5, and heterozygosity filtering was not performed.

After quality control, the ancestry stratified samples were combined and PRS were calculated using LDPred2.[3] Briefly, the LDPred2 tool leverages linkage disequilibrium information to provide a multivariate context for similarity across genetic variants. Accounting for this similarity, and not just removing closely linked variants, helps to improve prediction. We first convert bed files to the bigSNPR format as input for the PRS tool. We used the standard mean-zero imputation. In line with the PRS analyses conducted in the latest BD GWAS, we used a European LD reference for all PRS calculations.[4] PRS were calculated for BD, BDI and BDII using the latest and largest GWAS.[4,5] For BD we used summary statistics for both the European and the multiple ancestry GWAS. We resticted analyses to include only one relative (to the second degree) preferentially retaining BD cases over controls.
